# A predictive model for differentiating hemorrhagic fever with renal syndrome and scrub typhus in southwestern China

**DOI:** 10.64898/2026.03.02.26347402

**Authors:** Lihua Huang, Yunwei Zheng, Shumin Gu, Zihan Li, Fuxing Li, Wei Gu, Longhua Hu

## Abstract

**Background:** Both hemorrhagic fever with renal syndrome (HFRS) and scrub typhus (ST) are acute zoonotic infectious diseases. There is an overlap in their epidemiological characteristics and clinical manifestations, posing challenges for early differential diagnosis. This study aims to identify predictive factors for these two diseases to provide a basis for early diagnosis.

**Method/Findings:** A retrospective analysis was conducted on the clinical data of patients diagnosed with HFRS and ST at the First Affiliated Hospital of Dali University. Logistic regression analysis was employed to explore independent risk factors for the early differential diagnosis of these two diseases, and a nomogram model was constructed based on these risk factors. The performance of the model was evaluated using the area under the receiver operating characteristic curve (AUC). The nomogram was utilized to visually present the predictive variables. Decision curve analysis (DCA) was performed to assess the clinical utility of the model.

**Results:** A total of 235 patients each with HFRS and ST were included in this study. After adjusting for confounding factors, the results of multivariate logistic regression analysis revealed that sex (male) (adjusted odds ratio [ajOR]: 2.093, 95% confidence interval [CI]: 1.107 - 3.957, *P* = 0.018), positive proteinuria (ajOR: 4.937, 95% CI: 2.427 - 10.042, *P* < 0.001), creatinine (CREA) (ajOR: 1.009, 95% CI: 1.003 - 1.015, *P* = 0.005), heart rate (ajOR: 0.981, 95% CI: 0.966 - 0.997, *P* = 0.018), and conjunctival congestion (ajOR: 16.167, 95% CI: 5.326 - 49.072, *P* < 0.001) were independent risk factors for differentiating HFRS from ST. The AUC of the model constructed based on these five independent risk factors was 0.856.

**Conclusion:** Sex (male), positive proteinuria, elevated CREA, decreased heart rate, and conjunctival congestion are effective predictive factors.

## Introduction

In the field of global public health, zoonotic infectious diseases pose a severe threat to human health ^[1, 2]^. Hemorrhagic fever with renal syndrome (HFRS) is an acute zoonotic infectious disease of natural foci, caused by hantaviruses of the *Bunyaviridae* family, with rodents being the primary source of infection for this disease ^[3]^. According to epidemiological reports, HFRS is mainly prevalent in Europe and Asia. In the 20th century, China reported the highest number of cases globally ^[4]^, with a particular predominance in rural areas. The disease presents with a diverse range of clinical manifestations, including symptoms such as fever, headache, abdominal pain, hypotensive shock, and acute renal failure ^[5, 6]^. However, currently, there is a lack of effective antiviral drugs or vaccines, and the treatment mainly relies on supportive care ^[7]^. Our recent research has found that the case fatality rate of this disease is approximately 8.58% ^[8]^.

Scrub typhus (ST) is another zoonotic infectious disease, primarily caused by infection with *Orientia tsutsugamushi* ^[9, 10]^, with rodents being the main host ^[11]^. It has been reported that over one billion people worldwide are at risk of ST infection, with an annual incidence of up to one million cases ^[12, 13]^, posing a significant threat to public health. In China, the epidemic trend of ST shows a gradual spread from the southwest and south to the central and northern regions. However, provinces such as Guangdong and Yunnan remain high-incidence areas for this disease ^[14]^. Its typical clinical manifestations include fever, headache, eschar, etc. If not treated promptly, it can progress to multiple organ dysfunction and even death ^[13, 15, 16]^.

HFRS and ST not only share similar epidemiological distribution characteristics. Specifically, the warm and humid climate conditions and dense vegetation coverage in the Dali region of China provide an ideal habitat for rodents (the primary source of infection for HFRS) and trombiculid mites (the transmission vector of ST) ^[17]^, resulting in these two zoonotic infectious diseases posing a significant threat to population health in this area. They also exhibit overlaps in clinical features ^[18]^. The course of a typical HFRS patient can be divided into five characteristic clinical stages: febrile phase, hypotensive phase, oliguric phase, polyuric phase, and convalescent phase ^[6, 7]^, while patients with severe ST may develop multiple organ dysfunction, including shock, acute kidney injury, respiratory failure, etc. In clinical diagnosis and treatment practice, failure to accurately differentiate between these two diseases and formulate individualized treatment plans in the early stages of the diseases can not only delay treatment timing and affect clinical outcomes but also increase the economic burden on patients. Therefore, early and accurate differentiation between HFRS and ST is of great significance for optimizing clinical decision-making and improving patient prognosis. Hence, this study aims to explore the risk factors for differentiating between these two infectious diseases by integrating patients’ clinical features and laboratory indicators, providing a basis for clinical practice.

## Methods

### Ethical approval

This study adhered to the principles outlined in the Declaration of Helsinki and received approval from the Medical Ethics Committee of the First Affiliated Hospital of Dali University (No: 20251203001). All clinical and laboratory data were anonymized. Given that this study was retrospective in nature, the Medical Ethics Committee of the First Affiliated Hospital of Dali University granted a waiver of informed consent for the patients.

### Research design

This study conducted a retrospective analysis of the data from 235 patients diagnosed with HFRS and 235 patients diagnosed with ST at the First Affiliated Hospital of Dali University between January 1, 2015, and December 31, 2025. The diagnostic criteria for HFRS were based on the industry standard “Diagnostic Criteria for Epidemic Hemorrhagic Fever” issued by the Ministry of Health of the People’s Republic of China ^[19]^, with the specific criteria as follows: (1) Within 2 months prior to disease onset, the patient had a history of residing in an epidemic area or had contact with rodents and their excreta or secretions. (2) The patient presented with clinical manifestations such as fever, hemorrhage, renal injury, and hypotensive shock. (3) Laboratory indicators showed an elevated white blood cell count, a decreased platelet count, the presence of abnormal lymphocytes, positive urinary protein, and increased levels of serum creatinine and blood urea nitrogen. (4) The patient simultaneously met any one of the following conditions: ① Positive serum-specific IgM antibody; ② A four-fold or greater increase in the serum-specific IgG antibody titer during the convalescent phase compared to the acute phase; ③ Detection of hantavirus RNA in patient specimens; ④ Isolation of hantavirus from patient specimens.

The diagnostic criteria for ST were based on those outlined in the “Guidelines for the Prevention and Control of ST (2009 Edition)” issued by the Chinese Center for Disease Control and Prevention ^[20]^, with the specific criteria as follows: (1) Patients with a history of outdoor activities in an epidemic area within three weeks of disease onset and presenting with fever and eschar ulcer symptoms; or patients who, during the epidemic season, had no definite exposure history but presented with clinical manifestations such as fever, eschar ulcer, lymphadenopathy, and rash. (2) The patient simultaneously met any one of the following conditions: ① Positive Weil-Felix test; ② Positive indirect immune fluorescence antibody assay; ③ Positive polymerase chain reaction test; ④ Isolation and culture of *Orientia tsutsugamushi*.

The exclusion criteria for all patients were as follows: ① Age under 18 years; ② Comorbidities including malignant tumors, ongoing immunosuppressive drug therapy, hematologic disorders, chronic kidney diseases, etc.; ③ Missing clinical information exceeding 20%.

### Data collection

Relevant data within 24 hours after admission were extracted from the electronic medical record systems of patients, including: **Baseline Information**: sex, age, body temperature, breath, heart rate, systolic blood pressure, diastolic blood pressure, course of disease, length of hospital stay, pregnancy, intensive care unit (ICU) admission, receiving continuous renal replacement therapy (CRRT) and /or mechanical ventilation (MV), prognosis, place of residence, occupation. **Symptoms and Physical Signs**: Fever, headache, abdominal pain, diarrhea, dizzy, nausea, vomiting, cough, distress, chilly, shiver, dyspnea, fatigue, myalgia, eschar/rash, conjunctival hyperemia, etc. **Laboratory Indicators**: White blood cell (WBC), absolute neutrophil count (NEUT), absolute lymphocyte count (LYMPH), absolute monocyte count (MONO), red blood cell (RBC), hemoglobin (HGB), and platelet (PLT), total bilirubin (TBIL) level, direct bilirubin (DBIL), and indirect bilirubin (IBIL), alanine aminotransferase (ALT), aspartate aminotransferase (AST), alkaline phosphatase (ALP), and gamma-glutamyl transferase (GGT), albumin (ALB), total cholesterol (TC), triglyceride (TG), fasting blood glucose (FBG), lactate dehydrogenase (LDH), blood urea nitrogen (BUN), creatinine (CREA), and uric acid (UA), prothrombin time (PT), activated partial thromboplastin time (APTT), thrombin time (TT), and fibrinogen (FIB), serum potassium (K), serum sodium (Na), serum calcium (Ca), serum magnesium (Mg), serum phosphorus (P), procalcitonin (PCT), C-reactive protein (CRP), proteinuria, and hematuria. **Imaging Findings**: such as pulmonary infection, hydrothorax, etc.

In this study, proteinuria levels categorized as “-” to “++” were defined as negative, while those categorized as “+++” to “++++” were defined as positive.

### Statistical analysis

For categorical data, rates and constituent ratios were used for description, and the χ^2^ test or Fisher’s exact test was applied for inter-group comparisons. For continuous data, if the data conformed to a normal distribution, it was expressed as mean ± standard deviation, with the t-test employed for inter-group comparisons; if the data exhibited a non-normal distribution, it was described using the interquartile range, and the Mann-Whitney U test was utilized for inter-group comparisons. In the analysis of risk factors, variables with a *P* - value < 0.05 in univariate analysis were included in logistic regression analysis. All tests were conducted as two-tailed tests, with a significance level set at 0.05.

## Results

### Baseline characteristics

This study included a total of 235 cases each of patients with HFRS and ST, all of whom were laboratory-confirmed. HFRS cases predominantly occurred between April and June and between October and December, exhibiting a bimodal seasonal distribution (**Figure 1A**). Among the HFRS patients, there were 171 males and 64 females, with a male-to-female ratio of 2.67:1. The mean age was (44.85 ± 14.14) years. There were 198 rural patients and 37 urban patients, with the majority being farmers (154 cases, 65.53%) and forestry workers (67 cases, 28.51%).

**Figure 1A:**
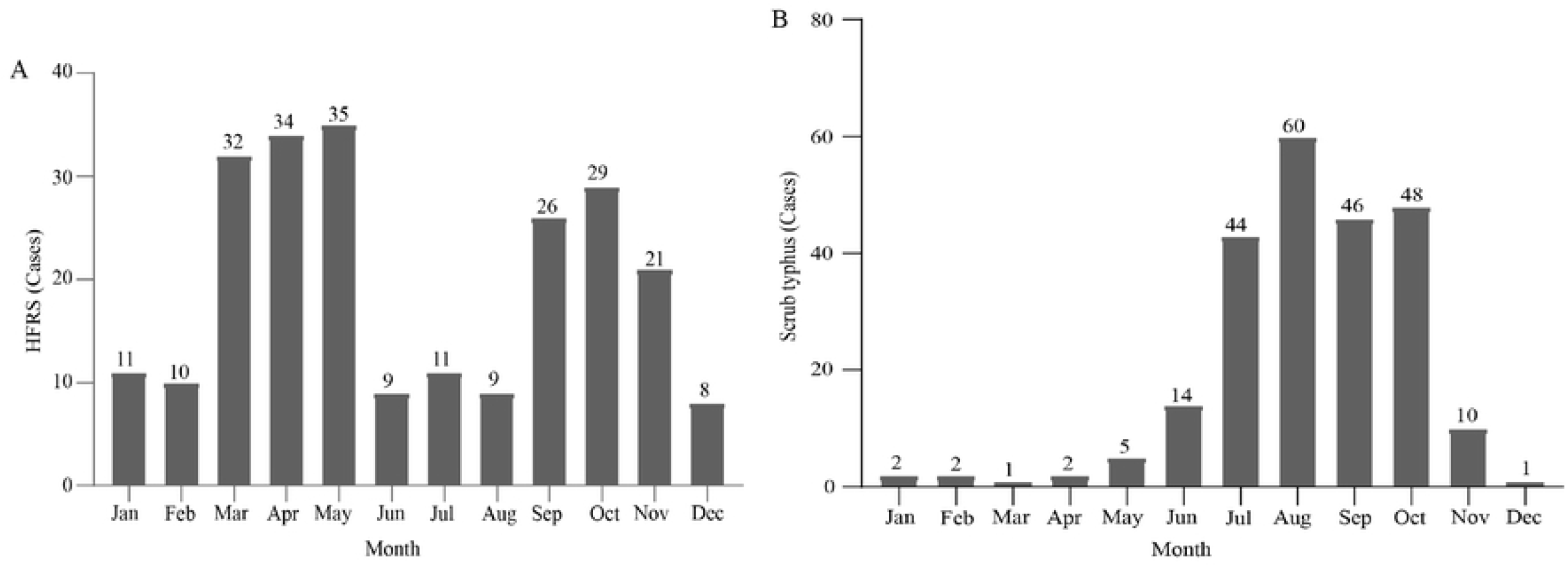
Bar chart showing the distribution of onset months in patients with HFRS. **Figure1B**: Bar chart illustrating the distribution of onset months in patients with ST.

ST cases were mainly concentrated from July to October, showing a high incidence trend in summer and autumn (**Figure 1B**). Among the ST patients, there were 96 males and 139 females, with a male-to-female ratio of 1:1.45. The mean age was (47.92 ± 14.27) years. There were 201 rural patients and 34 non-rural patients, with the majority being farmers (171 cases, 72.77%) and forestry workers (53 cases, 22.55%) (**Table 1**).

**Table 1.** Comparison of general conditions between patients with HFRS and ST.

| Variables | HFRS (n = 235) | ST (n = 235) | Statistic | P - value |
| --- | --- | --- | --- | --- |
| Body temperature (°C) | 39.00 (38.05, 39.50) | 39.00 (38.20, 39.50) | Z = -0.85 | 0.397 |
| Breath (Pulse beats per minute) | 20.00 (20.00, 20.00) | 20.00 (20.00, 20.00) | Z = -0.44 | 0.659 |
| Heart rate (Pulse beats per minute) | 86.34 ± 18.37 | 95.55 ± 17.04 | t = -5.64 | < .001 |
| Systolic blood pressure (mmHg) | 106.00 (97.00, 116.50) | 103.00 (95.00, 114.00) | Z = -1.34 | 0.181 |
| Diastolic blood pressure (mmHg) | 65.00 (60.00, 69.50) | 64.00 (58.00, 72.50) | Z = -0.34 | 0.730 |
| Age (year) | 44.85 ± 14.14 | 47.92 ± 14.27 | t = -2.35 | 0.019 |
| Sex, n (%) | | | $\chi^2 = 47.66$ | < .001 |
| Female | 63 (26.81) | 137 (58.30) |  |  |
| Male | 172 (73.191) | 98 (41.702) |  |  |
| Course of disease, (day) | 5.00 (4.00, 6.00) | 5.00 (4.00, 6.00) | Z = -1.89 | 0.058 |
| Length of hospital stay, (day) | 10.00 (8.00, 14.00) | 7.00 (5.00, 9.00) | Z = -9.59 | < .001 |
| Pregnancy | | | $\chi^2 = 0.29$ | 0.587 |
| No | 229 (97.45) | 227 (96.60) |  |  |
| Yes | 6 (2.55) | 8 (3.40) |  |  |
| ICU admission, n (%) | | | $\chi^2 = 4.96$ | 0.026 |
| No | 210 (89.36) | 223 (94.89) |  |  |
| Yes | 25 (10.64) | 12 (5.11) |  |  |
| Receiving CRRT and/or MV, n (%) | | | $\chi^2 = 13.16$ | < .001 |
| No | 186 (79.15) | 214 (91.06) |  |  |
| Yes | 49 (20.85) | 21 (8.94) |  |  |
| Prognosis, n (%) | | | $\chi^2 = 8.61$ | 0.003 |
| Survival | 216 (91.91) | 230 (97.87) |  |  |
| Death | 19 (8.09) | 5 (2.13) |  |  |
| Place of residence, n (%) | | | $\chi^2 = 0.15$ | 0.699 |
| Rural | 198 (84.26) | 201 (85.53) |  |  |
| Urban | 37 (15.74) | 34 (14.47) |  |  |
| Occupation, n (%) | | | $\chi^2 = 2.88$ | 0.237 |
| Farmer | 154 (65.53) | 171 (72.77) |  |  |
| Forestry worker | 67 (28.51) | 53 (22.55) |  |  |
| Other | 14 (5.96) | 11 (4.68) |  |  |

Notes: ICU: Intensive Care Unit; CRRT: Continuous Renal Replacement Therapy; MV: Mechanical Ventilation; HFRS: Hemorrhagic Fever with Renal Syndrome; ST: Scrub Typhus.

Compared with the ST group, the HFRS group showed statistically significant differences in the proportion of males, age, heart rate, mean length of hospital stay, proportion of patients admitted to the ICU, proportion of patients receiving Continuous Renal Replacement Therapy (CRRT) and/or invasive mechanical ventilation, proportion of pregnancy, and mortality rate (*P* < 0.05). However, no statistically significant differences were observed between the two groups in terms of place of residence and occupation (*P* > 0.05) (**Table 1**).

In terms of clinical manifestations, statistically significant differences were observed between the two groups in abdominal pain, diarrhea, nausea, vomiting, cough, fatigue, fever, eschar/rash, and conjunctival congestion (*P* < 0.05). However, no statistically significant differences were noted in dizzy, distress, chilly, shiver, dyspnea, myalgia, and headache between the two groups (**Table 2**). In terms of laboratory parameters, statistically significant differences were observed between the two groups in APTT, TT, TBIL, DBIL, IBIL, ALT, AST, ALP, BUN, CREA, UA, TG, WBC, NEUT, MONO, RBC, HGB, HCT, PLT, CK, CKMB, LDH, HBDH, proteinuria, and hematuria (*P* < 0.05). However, no statistically significant differences were noted in ALB, PCT, CRP, PT, FIB, GGT, K, Na, Ca, Mg, P, TC, LYMPH, pulmonary infection, and hydrothorax between the two groups (*P* > 0.05) (**Table 2**).

**Table 2.** Clinical manifestations and laboratory indicators of HFRS and ST.

| Variables | HFRS (n = 235) | ST (n = 235) | Statistic | P - value |
| --- | --- | --- | --- | --- |
| **Clinical manifestations** |  |  |  |  |
| Abdominal pain, n (%) | | | $\chi^2 = 23.76$ | < .001 |
| No | 195 (82.98) | 227 (96.60) |  |  |
| Yes | 40 (17.02) | 8 (3.40) |  |  |
| Diarrhea, n (%) | | | $\chi^2 = 17.56$ | < .001 |
| No | 213 (90.64) | 233 (99.15) |  |  |
| Yes | 22 (9.36) | 2 (0.85) |  |  |
| Dizzy, n (%) | | | $\chi^2 = 1.04$ | 0.309 |
| No | 225 (95.75) | 229 (97.45) |  |  |
| Yes | 10 (4.26) | 6 (2.55) |  |  |
| Nausea, n (%) | | | $\chi^2 = 8.33$ | 0.004 |
| No | 222 (94.47) | 233 (99.15) |  |  |
| Yes | 13 (5.53) | 2 (0.85) |  |  |
| Vomiting, n (%) | | | $\chi^2 = 14.23$ | < .001 |
| No | 214 (91.06) | 232 (98.72) |  |  |
| Yes | 21 (8.94) | 3 (1.28) |  |  |
| Cough, n (%) | | | $\chi^2 = 4.15$ | 0.042 |
| No | 219 (93.19) | 206 (87.66) |  |  |
| Yes | 16 (6.81) | 29 (12.34) |  |  |
| Distress, n (%) | | | $\chi^2 = 0.41$ | 0.523 |
| No | 229 (97.45) | 231 (98.30) |  |  |
| Yes | 6 (2.55) | 4 (1.70) |  |  |
| Chilly, n (%) | | | $\chi^2 = 1.78$ | 0.182 |
| No | 205 (87.23) | 214 (91.06) |  |  |
| Yes | 30 (12.77) | 21 (8.94) |  |  |
| Shiver, n (%) | | | $\chi^2 = 0.58$ | 0.446 |
| No | 233 (99.15) | 230 (97.87) |  |  |
| Yes | 2 (0.85) | 5 (2.13) |  |  |
| Dyspnea, n (%) | | | $\chi^2 = 2.65$ | 0.104 |
| No | 231 (98.30) | 225 (95.75) |  |  |
| Yes | 4 (1.70) | 10 (4.26) |  |  |
| Fatigue, n (%) | | | $\chi^2 = 5.03$ | 0.025 |
| No | 193 (82.13) | 210 (89.36) |  |  |
| Yes | 42 (17.87) | 25 (10.64) |  |  |
| Myalgia, n (%) | | | $\chi^2 = 1.28$ | 0.258 |
| No | 217 (92.34) | 223 (94.89) |  |  |
| Yes | 18 (7.66) | 12 (5.11) |  |  |
| Fever, n (%) | | | $\chi^2 = 11.54$ | < .001 |
| No | 24 (10.21) | 6 (2.55) |  |  |
| Yes | 211 (89.79) | 229 (97.45) |  |  |
| Headache, n (%) | | | $\chi^2 = 2.72$ | 0.099 |
| No | 178 (75.75) | 162 (68.95) |  |  |
| Yes | 57 (24.26) | 73 (31.06) |  |  |
| Conjunctival congestion | | | $\chi^2 = 54.01$ | < .001 |
| No | 175 (74.47) | 230 (97.87) |  |  |
| Yes | 60 (25.53) | 5 (2.13) |  |  |
| Eschar/rash, n (%) | | | $\chi^2 = 462.07$ | < .001 |
| No | 233 (99.15) | 0 (0.00) |  |  |
| Yes | 2 (0.85) | 235 (100.00) |  |  |
| **Laboratory indicators** |  |  |  |  |
| Procalcitonin ($\mu\text{g/L}$) | 1.05 (0.36, 1.60) | 0.71 (0.34, 1.38) | Z = -0.75 | 0.451 |
| C - reactive protein (mg/L) | 67.10 (25.00, 116.90) | 55.98 (23.45, 94.50) | Z = -1.67 | 0.096 |
| Prothrombin time (s) | 12.20 (11.30, 13.10) | 12.30 (11.50, 13.20) | Z = -1.12 | 0.263 |
| Activated partial thromboplastin time (s) | 33.00 (27.30, 42.45) | 31.40 (26.40, 37.15) | Z = -2.48 | 0.013 |
| Thrombin time (s) | 19.30 (17.50, 23.40) | 18.50 (17.10, 20.20) | Z = -3.41 | < .001 |
| Fibrinogen (g/L) | 2.97 (2.30, 3.82) | 3.22 (2.49, 3.92) | Z = -0.98 | 0.329 |
| Total bilirubin ($\mu\text{mol/L}$) | 13.90 (10.25, 19.00) | 11.50 (7.90, 15.60) | Z = -4.31 | < .001 |
| Direct bilirubin ($\mu\text{mol/L}$) | 4.80 (3.35, 7.00) | 3.800 (2.80, 5.60) | Z = -4.14 | < .001 |
| Indirect bilirubin ($\mu\text{mol/L}$) | 8.40 (5.35, 12.80) | 6.60 (4.60, 10.80) | Z = -2.98 | 0.003 |
| Alanine aminotransferase (U/L) | 76.00 (43.50, 151.50) | 63.00 (39.00, 104.00) | Z = -2.85 | 0.004 |
| Aspartate aminotransferase (U/L) | 89.00 (43.50, 186.00) | 73.00 (50.50, 118.00) | Z = -1.34 | 0.182 |
| Alkaline phosphatase (U/L) | 80.00 (62.50, 114.50) | 100.00 (72.50, 149.50) | Z = -4.10 | < .001 |
| Gamma-glutamyltransferase (U/L) | 89.50 (48.00, 141.50) | 90.00 (41.50, 194.50) | Z = -1.02 | 0.306 |
| Albumin (g/L) | 31.780 ± 5.88 | 32.38 ± 5.43 | t = -1.15 | 0.250 |
| Blood urea nitrogen (mmol/L) | 7.14 (4.84, 16.08) | 4.39 (3.41, 6.01) | Z = -8.93 | < .001 |
| Creatinine ($\mu\text{mol/L}$) | 110.00 (76.00, 264.50) | 67.00 (55.00, 80.00) | Z = -11.29 | < .001 |
| Uric acid ($\mu\text{mol/L}$) | 381.00 (257.50, 586.50) | 267.00 (224.50, 337.50) | Z = -7.28 | < .001 |
| Fasting blood glucose (mmol/L) | 5.72 (4.94, 7.34) | 5.62 (4.90, 6.66) | Z = -1.46 | 0.146 |
| Serum potassium (mmol/L) | 3.78 ± 0.54 | 3.71 ± 0.52 | t = 1.53 | 0.127 |
| Serum sodium (mmol/L) | 138.00 (135.00, 141.05) | 138.00 (135.85, 140.65) | Z = -0.39 | 0.699 |
| Serum calcium (mmol/L) | 2.02 (1.90, 2.15) | 2.05 (1.95, 2.14) | Z = -1.45 | 0.147 |
| Serum magnesium (mmol/L) | 0.87 (0.79, 0.98) | 0.86 (0.80, 0.95) | Z = -0.77 | 0.439 |
| Serum inorganic phosphorus (mmol/L) | 1.03 (0.84, 1.38) | 1.01 (0.87, 1.20) | Z = -1.16 | 0.246 |
| Total cholesterol (mmol/L) | 3.19 (2.63, 3.85) | 3.22 (2.70, 3.88) | Z = -0.73 | 0.465 |
| Triglyceride (mmol/L) | 2.30 (1.73, 3.26) | 2.10 (1.55, 2.71) | Z = -2.90 | 0.004 |
| White blood cell $\times 10^9/L$ | 8.24 (5.70, 11.26) | 6.87 (5.10, 9.31) | Z = -3.59 | < .001 |
| Absolute neutrophil count $\times 10^9/L$ | 4.74 (3.07, 6.79) | 4.17 (2.81, 5.75) | Z = -2.60 | 0.009 |
| Absolute lymphocyte count $\times 10^9/L$ | 1.69 (1.05, 2.79) | 1.69 (1.02, 2.93) | Z = -0.05 | 0.963 |
| Absolute monocyte count $\times 10^9/L$ | 0.81 (0.52, 1.24) | 0.50 (0.34, 0.74) | Z = -6.67 | < .001 |
| Red blood cell $\times 10^{12}/L$ | 4.76 (4.25, 5.28) | 4.33 (3.93, 4.70) | Z = -6.59 | < .001 |
| Hemoglobin (g/L) | 141.00 (125.00, 155.50) | 130.00 (116.00, 140.00) | Z = -6.07 | < .001 |
| Hematocrit (%) | 42.50 (37.75, 46.85) | 38.50 (34.85, 41.85) | Z = -6.76 | < .001 |
| Platelet $\times 10^9/L$ | 70.00 (44.00, 144.50) | 128.00 (90.00, 196.50) | Z = -7.24 | < .001 |
| Atypical lymphocytes, n (%) | | | $\chi^2 = 0.04$ | 0.841 |
| No | 162 (68.94) | 164 (69.79) |  |  |
| Yes | 73 (31.06) | 71 (30.21) |  |  |
| Creatine kinase (U/L) | 69.00 (40.00, 146.50) | 60.00 (38.00, 110.00) | Z = -2.20 | 0.028 |
| Creatine kinase - MB (U/L) | 15.00 (8.100, 24.00) | 12.00 (9.00, 17.00) | Z = -3.84 | < .001 |
| Lactate dehydrogenase (U/L) | 444.00 (317.00, 693.50) | 400.00 (295.00, 498.50) | Z = -3.48 | < .001 |
| α-Hydroxybutyrate dehydrogenase (U/L) | 320.00 (242.00, 472.50) | 296.00 (223.50, 375.50) | Z = -2.84 | 0.005 |
| Proteinuria, n (%) | | | $\chi^2 = 39.30$ | < .001 |
| No | 159 (67.66) | 214 (91.06) |  |  |
| Yes | 76 (32.34) | 21 (8.94) |  |  |
| Hematuria, n (%) | | | $\chi^2 = 4.14$ | 0.042 |
| No | 99 (42.13) | 121 (51.49) |  |  |
| Yes | 136 (57.87) | 114 (48.51) |  |  |
| Pulmonary infection, n (%) | | | $\chi^2 = 0.94$ | 0.332 |
| No | 159 (67.66) | 149 (63.40) |  |  |
| Yes | 76 (32.34) | 86 (36.60) |  |  |
| Hydrothorax, n (%) | | | $\chi^2 = 1.20$ | 0.274 |
| No | 156 (66.38) | 167 (71.06) |  |  |
| Yes | 79 (33.62) | 68 (28.94) |  |  |

Notes: HFRS: Hemorrhagic Fever with Renal Syndrome; ST: Scrub Typhus.

### Determinant associated with the differential diagnosis of HFRS and ST

Upon incorporating the indicators with statistically significant differences (*P* < 0.05) between HFRS and ST into univariate logistic regression analysis, it was found that age, heart rate, course of disease, APTT, TT, TBIL, IBIL, ALT, ALP, UA, CREA, UA, TG, WBC, NEUT, MONO, RBC, HGB, HCT, PLT, CK, CK-MB, LDH, HBDH, Sex, abdominal pain, diarrhea, nausea, vomiting, cough, fatigue, fever, conjunctival congestion, proteinuria, and hematuria were influencing factors for differentiating HFRS from ST (**Table 3**).

**Table 3.**
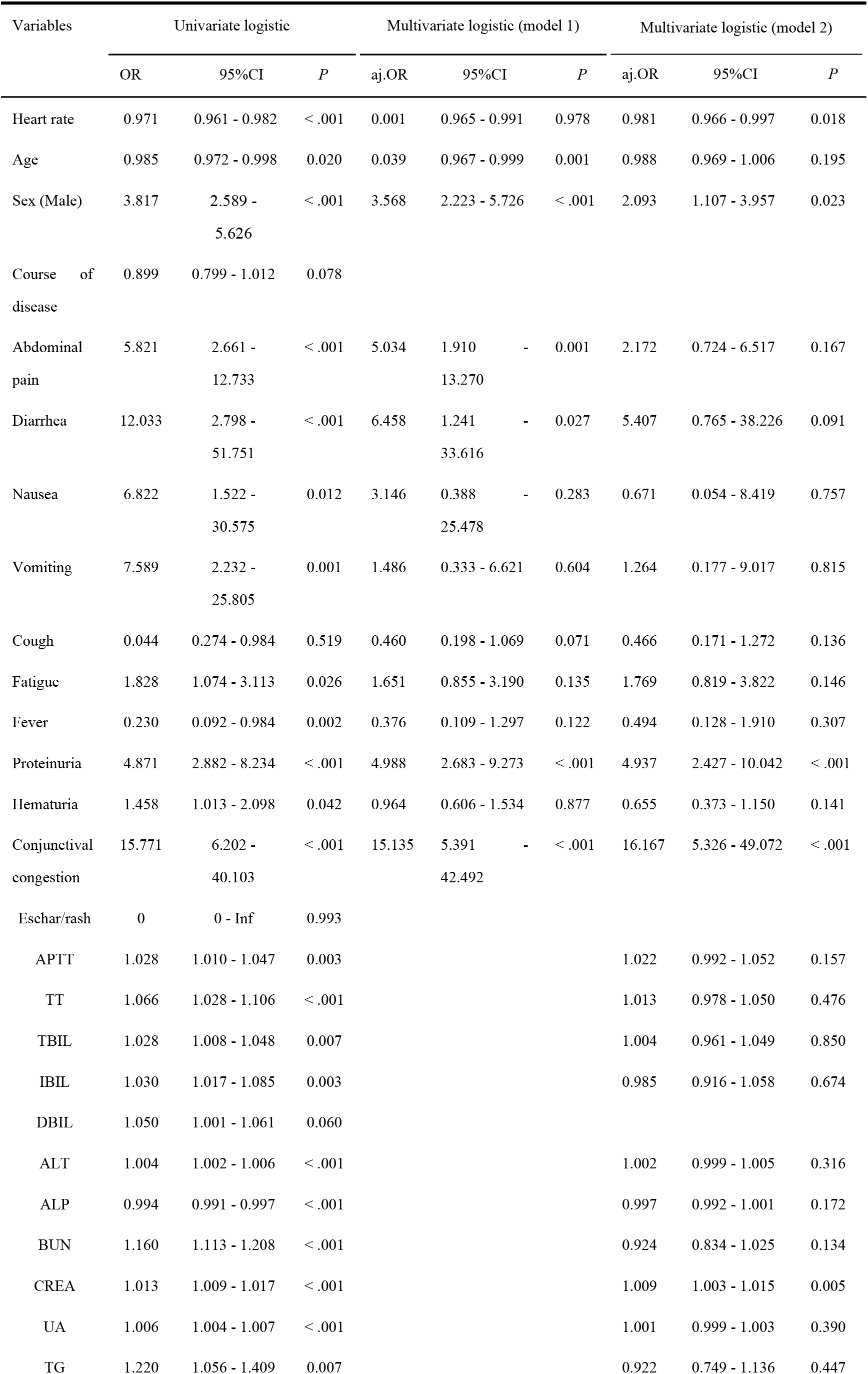
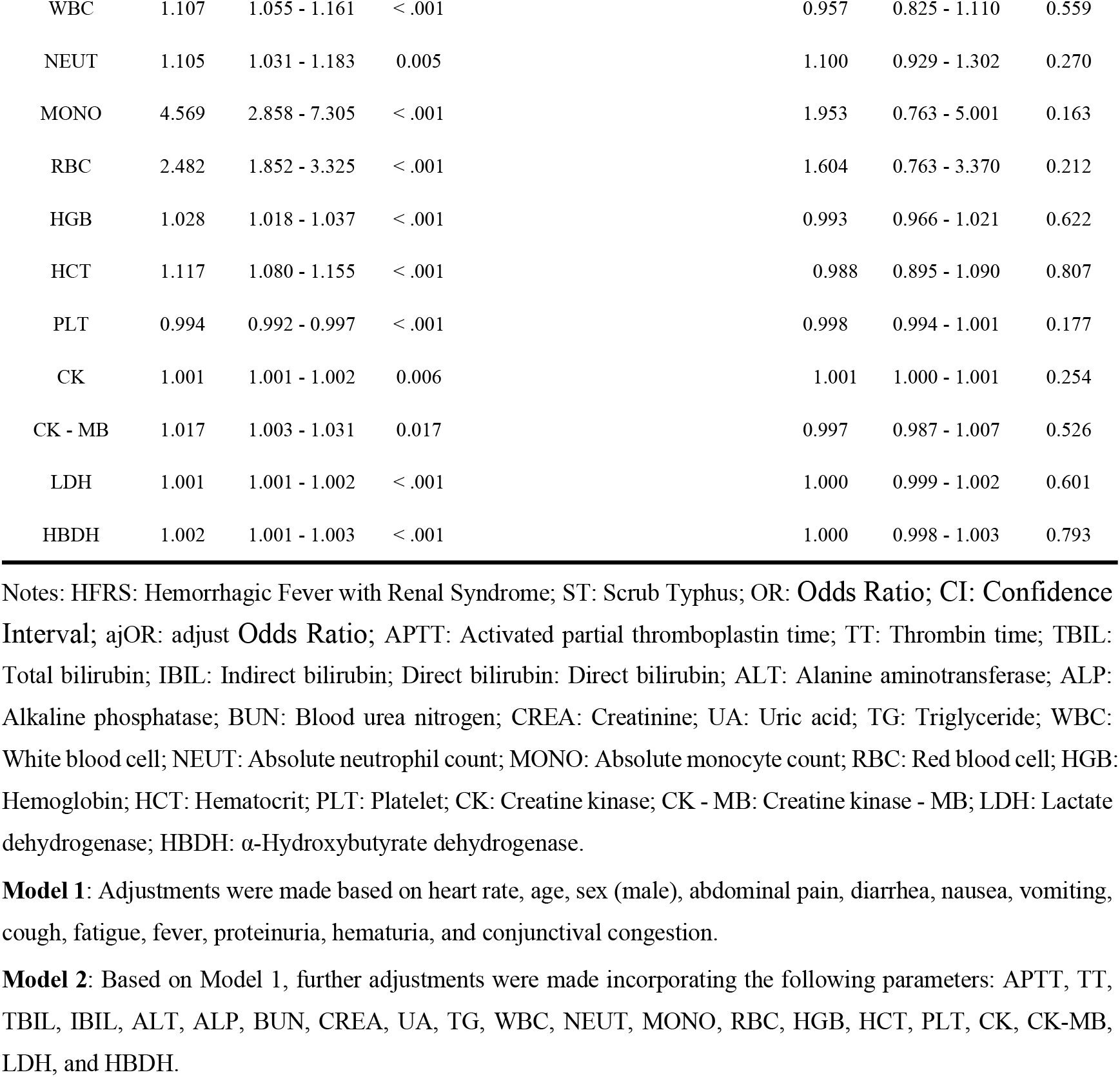
Univariate and multivariate logistic regression analyses were performed separately on predictive variables for HFRS and ST.

Subsequently, after conducting multivariate logistic regression analysis on the indicators that were statistically significant in the univariate logistic regression analysis and adjusting for confounding factors, it was found that sex (male) (adjusted odds ratio [ajOR]: 2.093, 95% confidence interval [CI]: 1.107 - 3.957, *P* = 0.018), positive proteinuria (ajOR: 4.937, 95% CI: 2.427 - 10.042, *P* < 0.001), CREA (ajOR: 1.009, 95% CI: 1.003 - 1.015, *P* = 0.005), heart rate (ajOR: 0.981, 95% CI: 0.966 - 0.997, *P* = 0.018), and conjunctival congestion (ajOR: 16.167, 95% CI: 5.326 - 49.072, *P* < 0.001) were independent risk factors for early differentiation between HFRS and ST (**Table 3**).

### Construction and evaluation of a differential diagnosis model for distinguishing HFRS from ST

A nomogram model was constructed for the early differentiation between HFRS and ST based on sex (male), positive proteinuria, CREA, heart rate, and conjunctival congestion (**Figure 2**). The receiver operating characteristic (ROC) curve was employed to evaluate the predictive performance of the differential diagnosis model, revealing an area under the curve (AUC) of 0.856 (**Figure 3**), indicative of favorable predictive capability. Additionally, the calibration curve of the nomogram-based differentiation model (**Figure 4A**) demonstrated close alignment between predicted values and actual incidence probabilities, reflecting good consistency and suggesting high accuracy of the model in differentiating HFRS from ST. Finally, the decision curve analysis (DCA) curve indicated that the differential diagnosis model provided a high clinical net benefit across a threshold probability range from 10% to 95% (**Figure 4B**).

**Figure 2.**
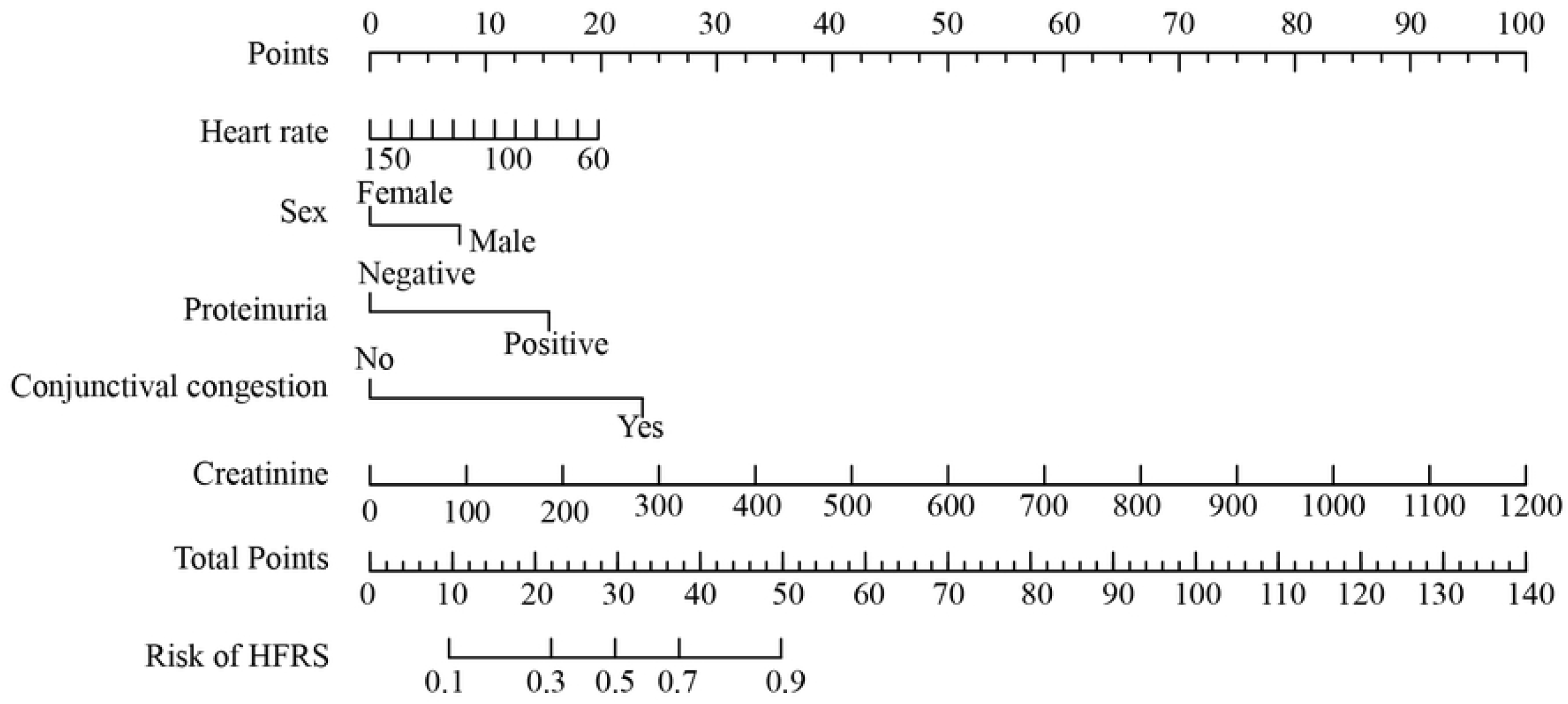
The nomogram of the model for differentiating HFRS from ST.

**Figure 3.**
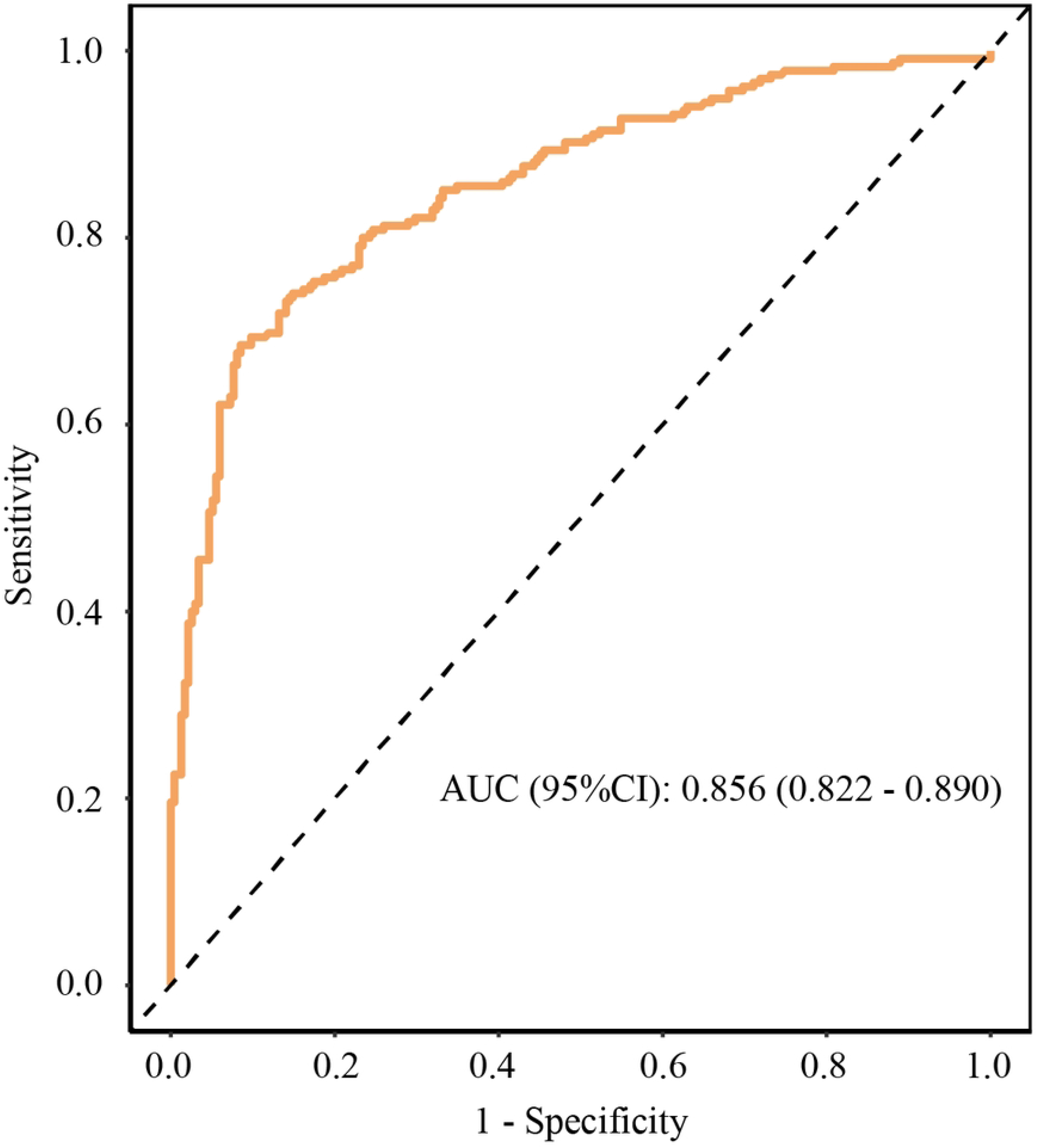
The Receiver Operating Characteristic Curve of the model for differentiating HFRS from ST.

**Figure 4A.**
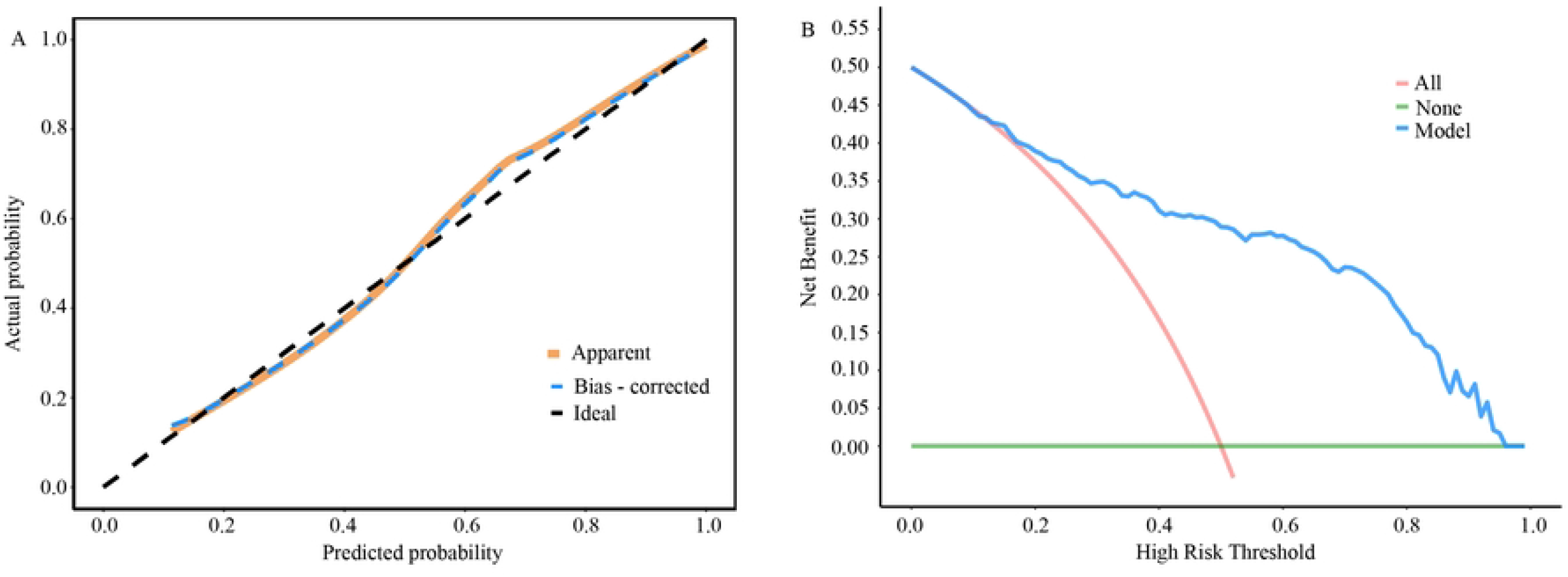
The calibration curve of the model for differentiating Hemorrhagic Fever with Renal Syndrome from scrub typhus. **Figure 4B**: The Decision Curve Analysis curve of the model for differentiating HFRS from ST.

## Discussion

Both HFRS and ST are fever-associated diseases that are prone to being overlooked. Despite their distinct pathogenic mechanisms, these two conditions exhibit overlapping epidemiological characteristics (such as geographical distribution and peak incidence seasons) and clinical manifestations (including fever, headache, and generalized body aches), posing challenges for early clinical differential diagnosis. Through a comparative analysis of the clinical features and laboratory indicators of HFRS and ST, this study constructed a diagnostic model for the early differentiation between these two diseases, with the following three key findings: (1) HFRS demonstrates a bimodal seasonal pattern with peaks in spring and autumn, whereas scrub typhus predominantly occurs in summer and autumn. (2) Sex (male), positive proteinuria, CREA, heart rate, and conjunctival congestion are independent risk factors for differentiating between these two diseases. (3) The nomogram model constructed based on the aforementioned risk factors exhibits favorable predictive performance and clinical utility, providing clinicians with a practical tool for the early differential diagnosis of these two diseases.

In this study, HFRS predominantly occurred in spring and autumn ^[21]^, whereas ST mainly presented in summer and autumn ^[22]^, with overlapping epidemic seasons. This pattern is attributable to the study site being located in the southern extension of the Hengduan Mountains in western Yunnan, characterized by significant elevation gradients and a subtropical monsoon climate. The natural environment during spring, summer, and autumn is conducive to the proliferation of trombiculid mites and rodents, particularly in spring during sowing and autumn during harvesting ^[23]^, thereby increasing the risk of human infection.

The proportion of males in the HFRS group was higher than that in the ST group. Further research revealed that sex (male) is one of the risk factors for differentiating between these two diseases. Previous studies have found that among HFRS patients, the prevalence rate in males is significantly higher than that in females ^[24]^, with a male-to-female patient ratio as high as 2.6:1 ^[3]^, suggesting that males are a high-risk population for HFRS onset. Recently, a cross-sectional study conducted by South Korean scholars on the clinical and laboratory characteristics of patients with HFRS and ST ^[18]^further confirmed that the incidence of HFRS is significantly higher in males than in females. Conversely, the proportion of females among ST patients is relatively higher ^[25]^. This may be closely related to activity patterns and occupational exposure factors in the population. Males are more likely to be engaged in agricultural and forestry work, often with longer working hours and a wider geographical range of activities, which increases their opportunities for contact with hantavirus-carrying rodents and their excreta, thereby elevating the risk of hantavirus infection. For ST, the relatively higher proportion of females may be associated with their roles in family life and specific activity settings. Females often take on more responsibilities in childcare and household chores within the family, potentially leading to more frequent contact with grassy and shrubby areas around the home, which are common habitats for trombiculid mites (the vectors of ST), thus increasing the risk of ST infection. Additionally, some females may be more inclined to participate in gardening activities, which also increases their exposure to environments where trombiculid mites thrive.

Although the pathological mechanisms of HFRS and ST have not been fully elucidated, increased endothelial permeability mediated by immune responses is considered the primary mechanism for both conditions ^[3, 26]^. In HFRS, the predominantly affected blood vessels are the renal medullary capillaries ^[27]^, manifesting clinically as acute kidney injury (AKI), elevated serum creatinine levels, and significant proteinuria. However, ST can involve systemic small blood vessels and is associated with loss of vascular endothelial integrity ^[28]^. While proteinuria and renal injury may also occur in ST, previous studies have reported that the incidence of ST-related AKI ranges from 12% to 41% ^[29, 30]^, with a generally milder degree of renal impairment compared to that observed in HFRS patients.

Hantavirus infection can activate immune cells ^[31]^, trigger the release of pro-inflammatory cytokines, simultaneously activate platelets and endothelial cells, and disrupt coagulation system function, thereby laying the groundwork for the occurrence of cardiovascular events ^[32]^. Our study found that patients with HFRS exhibited slower heart rates. Previous studies have suggested that 35%-80% of patients infected with hantavirus develop bradycardia ^[33, 34]^, with one study reporting that 80% of hantavirus-infected patients experience relative bradycardia. Additionally, compared to ST, conjunctival hyperemia was observed in a higher proportion of HFRS patients, which may be associated with capillary dilation. Finally, we constructed a model for differentiating between these two diseases based on five risk factors and visualized it using a nomogram. The AUC of the differentiation model was 0.856, indicating good predictive performance. The DCA demonstrated that the model exhibited favorable clinical applicability. Additionally, our study represents the first attempt to develop a nomogram model for the early differentiation between HFRS and ST. However, our study still has certain limitations. Firstly, this study only utilized single-center data with a relatively small sample size, which to some extent weakened the statistical power of the multivariate logistic regression analysis. Secondly, the lack of internal and external validation has somewhat restricted the applicability of the study results. Thirdly, due to the fact that indicators such as different subtypes of hantavirus, genotypes of ST patients, troponin, and quantitative urine protein are not routine clinical detection items and are difficult to obtain, this study failed to analyze these valuable indicators. Fourthly, this study did not conduct matched analysis on the severity of these two diseases. In view of the aforementioned limitations, we will make improvements and refinements in subsequent studies.

## Conclusion

This study constructed a nomogram prediction model for the early differentiation between HFRS and ST based on five variables: sex (male), positive proteinuria, CREA, heart rate, and conjunctival congestion. The model demonstrates good discriminatory ability and clinical decision-making value, aiding primary healthcare institutions in the early diagnosis and prompt treatment of these two diseases.

## Data Availability

All data generated or analysed during this study are included in this published article (and its supplementary information files

## Acknowledgments

We sincerely express our gratitude to all the medical staff who participated in this study, with special thanks extended to the Department of Infectious Diseases at the First Affiliated Hospital of Dali University for their support.

## Authors contributions

**Lihua Huang**: Conceptualization, Data curation, Formal analysis, Methodology, Resources, Writing – original draft; **Yunwei Zheng**: Data curation; **Shumin Gu**: Data curation; **Zihan L**i: Data curation; **Fuxing Li**: Writing – review & editing; **Wei Gu**: Formal analysis, Writing – review & editing; **Longhua Hu**: Conceptualization, Funding acquisition, Supervision.

## References

[1] Di Bari C, Venkateswaran N, Fastl C, Gabriel S, Grace D, Havelaar AH, et al. The global burden of neglected zoonotic diseases: Current state of evidence [J]. One Health, 2023, 17: 100595. doi:10.1016/j.onehlt.2023.100595

[2] Zhang Y, Geng M, Shi Y, Jin B, Xiong Q, Zhou S, et al. Epidemiological Characteristics and Trends of Zoonotic Diseases in China from 2015 to 2022 [J]. Trop Med Inf ect Dis, 2025, 10(6):159. doi:10.3390/tropicalmed10060159

[3] Vial PA, Ferres M, Vial C, Klingstrom J, Ahlm C, Lopez R, et al. Hantavirus in humans: a review of clinical aspects and management [J]. Lancet Infect Dis, 2023, 23(9): e371–e82. doi:10.1016/S1473-3099(23)00128-7

[4] Vaheri A, Henttonen H, Voutilainen L, Mustonen J, Sironen T, Vapalahti O. Hantavirus infections in Europe and their impact on public health [J]. Rev Med Virol, 2013, 23(1): 35–49. doi:10.1002/rmv.1722

[5] Wang Y, Zhang C, Gao J, Chen Z, Liu Z, Huang J, et al. Spatiotemporal trends of hemorrhagic fever with renal syndrome (HFRS) in China under climate variation [J]. Proc Natl Acad Sci U S A, 2024, 121(4): e2312556121. doi:10.1073/pnas.2312556121

[6] Jiang H, Du H, Wang LM, Wang PZ, Bai XF. Hemorrhagic Fever with Renal Syndrome: Pathogenesis and Clinical Picture [J]. Front Cell Infect Microbiol, 2016, 6: 1. doi: 10.3389/fcimb.2016.00001

[7] Lu W, Kuang L, Hu Y, Shi J, Li Q, Tian W. Epidemiological and clinical characteristics of death from hemorrhagic fever with renal syndrome: a meta-analysis [J]. Front Microbiol, 2024, 15: 1329683. doi:10.3389/fmicb.2024.1329683

[8] Huang L, Yan Q, Gao XM, Gu W. Risk Factors for 30-Day Prognosis of Hemorrhagic Fever with Renal Syndrome in the Dali Region, China [J]. Infect Drug Resist, 2025, 18: 6007–17. doi:10.2147/IDR.S549717

[9] Varghese GM, Dayanand D, Gunasekaran K, Kundu D, Wyawahare M, Sharma N, et al. Intravenous Doxycycline, Azithromycin, or Both for Severe Scrub Typhus [J]. N E ngl J Med, 2023, 388(9): 792–803. doi:10.1056/NEJMoa2208449

[10] Nie Y, Yang S, Yao Q, Liu X, Zhang B, Lu Y, et al. Epidemiological characteristics and influencing factors of scrub typhus in Jiangxi Province [J]. Parasit Vectors, 2025, 18(1): 260. doi:10.1186/s13071-025-06908-7

[11] Yang S, Yang S, Xie Y, Duan W, Cui Y, Peng A, et al. The Association Between Environmental Factors and Scrub Typhus: A Review [J]. Trop Med Infect Dis, 2025, 10 (6):151. doi:10.3390/tropicalmed10060151

[12] Jiang J, Richards AL. Scrub Typhus: No Longer Restricted to the Tsutsugamushi Triangle [J]. Trop Med Infect Dis, 2018, 3(1):11. doi:10.3390/tropicalmed3010011

[13] Jain HK, Das A, Dixit S, Kaur H, Pati S, Ranjit M, et al. Development and implementation of a strategy for early diagnosis and management of scrub typhus: an emerging public health threat [J]. Front Public Health, 2024, 12: 1347183. doi:10.3389/fpubh.2024.1347183

[14] Peng PY, Xu L, Sun JQ, Yan TL, Li ZL, Duan HY, et al. Epidemiologic features and potential year of life lost of scrub typhus in China: A nationwide surveillance analysis (2006-2023) [J]. PLoS Negl Trop Dis, 2025, 19(10): e0013666. doi:10.1371/journal.pntd.0013666

[15] Xu G, Walker DH, Jupiter D, Melby PC, Arcari CM. A review of the global epidemiology of scrub typhus [J]. PLoS Negl Trop Dis, 2017, 11(11): e0006062. doi:10.1371/journal.pntd.0006062

[16] Bhandari M, Singh RK, Laishevtcev A, Mohapatra TM, Nigam M, Mori E, et al. Revisiting scrub typhus: A neglected tropical disease [J]. Comp Immunol Microbiol Infect Dis, 2022, 90-91: 101888. doi:10.1016/j.cimid.2022.101888

[17] Wu YC, Qian Q, Soares Magalhaes RJ, Han ZH, Hu WB, Haque U, et al. Spatiotem poral Dynamics of Scrub Typhus Transmission in Mainland China, 2006-2014 [J]. PLo S Negl Trop Dis, 2016, 10(8): e0004875. doi:10.1371/journal.pntd.0004875

[18] Dorji T, Kim CM, Lee YM, Seo JW, Kim DY, Yun NR, et al. Clinical and laboratory factors associated with scrub typhus and hemorrhagic fever with renal syndrome in Southwestern Korea: A cross-sectional study [J]. PLoS Negl Trop Dis, 2025, 19(11): e0013709. doi:10.1371/journal.pntd.0013709

[19] Department of Health of the People’s Republic of China. Diagnostic criteria for epidemic hemorrhagic fever: WS 278-2008. Beijing: China Standards Press; 2008:1–11. Available from: http://www.nhc.gov.cn/wjw/s9491/200802/39043.shtml

[20] National guideline of scrub typhus control and prevention (in Chinese). Chinese Center for Disease Control and Prevention. 2009. http://www.chinacdc.cn/tzgg/200901/t20090105_40316.html

[21] Huang H, Fu M, Han P, Yin H, Yang Z, Kong Y, et al. Clinical and Molecular Epidemiology of Hemorrhagic Fever with Renal Syndrome Caused by Orthohantaviruses in Xiangyun County, Dali Prefecture, Yunnan Province, China [J]. Vaccines (Basel), 2023, 11(9). doi:10.3390/vaccines11091477

[22] Li Z, Deng S, Ma T, Hao J, Wang H, Han X, et al. Retrospective analysis of spatiotemporal variation of scrub typhus in Yunnan Province, 2006-2022 [J]. PLoS Negl Trop Dis, 2024, 18(12): e0012654. doi:10.1371/journal.pntd.0012654

[23] Lyu Y, Tian L, Zhang L, Dou X, Wang X, Li W, et al. A case-control study of risk factors associated with scrub typhus infection in Beijing, China [J]. PLoS One, 2013, 8(5): e63668. doi:10.1371/journal.pone.0063668

[24] Klein SL, Marks MA, Li W, Glass GE, Fang LQ, Ma JQ, et al. Sex differences in th e incidence and case fatality rates from hemorrhagic fever with renal syndrome in China, 2004-2008 [J]. Clin Infect Dis, 2011, 52(12): 1414–21. doi:10.1093/cid/cir232

[25] Devamani C, Alexander N, Chandramohan D, Stenos J, Cameron M, Abhilash KPP, et al. Incidence of Scrub Typhus in Rural South India [J]. N Engl J Med, 2025, 392(11): 1089–99. doi:10.1056/NEJMoa2408645

[26] Diaz FE, Abarca K, Kalergis AM. An Update on Host-Pathogen Interplay and Modulation of Immune Responses during Orientia tsutsugamushi Infection [J]. Clin Microbiol Rev, 2018, 31(2). doi:10.1128/CMR.00076-17

[27] Noack D, Goeijenbier M, Reusken C, Koopmans MPG, Rockx BHG. Orthohantavirus Pathogenesis and Cell Tropism [J]. Front Cell Infect Microbiol, 2020, 10: 399. doi:10.3389/fcimb.2020.00399

[28] Shelite TR, Saito TB, Mendell NL, Gong B, Xu G, Soong L, et al. Hematogenously disseminated Orientia tsutsugamushi-infected murine model of scrub typhus [corrected] [J]. PLoS Negl Trop Dis, 2014, 8(7): e2966. doi:10.1371/journal.pntd.0002966

[29] Basu G, Chrispal A, Boorugu H, Gopinath KG, Chandy S, Prakash JA, et al. Acute kidney injury in tropical acute febrile illness in a tertiary care centre--RIFLE criteria validation [J]. Nephrol Dial Transplant, 2011, 26(2): 524–31. doi:10.1093/ndt/gfq477

[30] Sun IO, Kim MC, Park JW, Yang MA, Lee CB, Yoon HJ, et al. Clinical characteristics of acute kidney injury in patients with scrub typhus--RIFLE criteria validation [J]. J Infect Chemother, 2014, 20(2): 93–6. doi:10.1016/j.jiac.2013.08.007

[31] Garcia M, Carrasco Garcia A, Weigel W, Christ W, Lira-Junior R, Wirth L, et al. Inn ate lymphoid cells are activated in HFRS, and their function can be modulated by hantavirus-induced type I interferons [J]. PLoS Pathog, 2024, 20(7): e1012390. doi:10.1371/journal.ppat.1012390

[32] Connolly-Andersen AM, Hammargren E, Whitaker H, Eliasson M, Holmgren L, Klingstrom J, et al. Increased risk of acute myocardial infarction and stroke during hemorrhagic fever with renal syndrome: a self-controlled case series study [J]. Circulation, 2014, 129(12): 1295–302. doi:10.1161/CIRCULATIONAHA.113.001870

[33] Pal E, Korva M, Resman Rus K, Kejzar N, Bogovic P, Kurent A, et al. Sequential assessment of clinical and laboratory parameters in patients with hemorrhagic fever with renal syndrome [J]. PLoS One, 2018, 13(5): e0197661. doi:10.1371/journal.pone.0197661

[34] Kitterer D, Greulich S, Grun S, Segerer S, Mustonen J, Alscher MD, et al. Electrocardiographic abnormalities and relative bradycardia in patients with hantavirus-induced nephropathia epidemica [J]. Eur J Intern Med, 2016, 33: 67–73. doi:10.1016/j.ejim.2016.06.001

